# Energy dense nutritional supplements improve weight gain among malnourished adults with drug-sensitive pulmonary tuberculosis: an open-label randomized controlled trial in Faridabad, India

**DOI:** 10.1101/2025.07.15.25331513

**Authors:** Rakesh Kumar, Pranay Sinha, Anand Krishnan, Manjula Singh, Archna Singh, Randeep Guleria, Urvashi B Singh

## Abstract

This open-label randomized controlled trial in India assessed the impact of a peanut-based energy-dense nutritional supplement (EDNS) on weight gain among undernourished adults with drug-susceptible pulmonary tuberculosis in India. EDNS significantly improved weight gain compared to standard care, offering a scalable solution for targeted nutritional support.

## Introduction

India carries a quarter of the global TB burden.^1^ Undernutrition, which is associated with increased disease severity and unfavorable outcomes, is the leading driver for TB in India.^2^ Poor weight gain is associated with increased mortality whereas gaining at least 5% of baseline body weight during the intensive phase is associated with reduced mortality.^3,4^

There is a paucity of randomized studies that have reported the impact of macronutrient support on weight gain in persons with tuberculosis (PWTB). Indeed, the 2016 COCHRANE review reported only included five underpowered studies that assessed the impact of macronutrient support on weight gain.^2,5^ Although PWTB in the RATIONS study showed meaningful weight gain, the lack of a control group makes it difficult to discern the impact of nutritional support. Extrapolating weight gain data from studies of general populations is unwise, as TB induces profound metabolic alterations.^2^

Here, we present data from an open-label, randomized-controlled trial of an energy-dense nutritional supplement (EDNS) to understand its impact on weight gain during TB treatment.

## Methods

### Study Design and Setting

We conducted this open-label, two-arm randomized controlled trial between 2020 and 2023 in two tuberculosis (TB) units in Faridabad district, Haryana, India. Ethics committee of All India Institute of Medical Sciences (AIIMS), New Delhi, approved the study (IEC-110/09.03.2018, RP-23/2018, OP-7/2.8.2019). The trial was registered with Central Trial Registry of India (CTRI/2019/12/022574).

### Participants

We enrolled adults (≥18 years) with microbiologically confirmed, drug-sensitive pulmonary TB and mild to moderate undernutrition (BMI 14–18.5 kg/m^2^) receiving treatment under the National TB Elimination Programme (NTEP). We excluded individuals with peanut allergies, pregnancy or lactation, prior diabetes diagnosis or fasting glucose/HbA1c abnormalities, HIV seropositivity, renal or hepatic impairment, or poor general condition. Participants provided≥ written consent, and the.

### Randomization and Blinding

We randomized participants to either the intervention or control arm using a computer-generated sequence and ensured allocation concealment with sequentially numbered, opaque, sealed envelopes. Blinding was not feasible for participants or investigators.

### Intervention

Participants in the intervention arm received a previously described EDNS weekly alongside standard dietary advice. Each 92 g EDNS sachet provided 500 kcal, 13 g protein, 29 g fat, and micronutrients.^6^ They consumed two sachets daily for six months or until reaching a BMI of 21 kg/m^2^. The control group received only standard dietary advice. Both groups received standard TB treatment.

### Outcome Assessments

The primary outcomes were 5% weight gain from baseline at two months and 10% at treatment completion. We weighed participants monthly. Secondary outcomes included EDNS acceptability (assessed via a structured questionnaire on taste, smell, consistency, color, and ease of use after one month), adherence (measured by counting returned empty sachets), adverse effects (recorded biweekly), and hemoglobin improvement.

### Data Collection and Management

We collected data electronically using the Open Data Kit (ODK) platform on Android devices and securely stored it on a local MySQL server.

### Statistical Analysis

We used box plots to compare percent weight gain in the control and intervention groups over six months. Multivariable logistic regression estimated the adjusted odds ratio for achieving 5% weight gain at month 2 and 10% at treatment completion. *A priori*, we adjusted for age, sex, asset quintile, and baseline BMI and included covariates with p-values <0.20 in baseline characteristics or univariable regression. We presented acceptability and adverse effect frequencies as proportions.

## Results

### Recruitment

We randomly allocated 335 participants to the intervention or control arm, with 171 in the intervention arm and 164 in the control arm. We subsequently excluded six participants (four intervention, two control) upon detecting multidrug resistance (Supplementary Figure 1).

### Demographics

Mean age of participants was not significantly different in the intervention and control arms (33.1 ± 13.4 vs. 33.4 ± 14.7 years, p = 0.802). There was a higher proportion of men in the control arm (65.9% vs. 59.2%). The mean BMI of participants (p= 0.25) and distribution across wealth quintiles (p= 0.701) were similar between arms. (Supplementary table 1)

### Weight gain trends

Participants in the intervention group gained a higher percentage of their baseline body weight in both the intensive phase and the continuation phase. (Figure 1).At the end of the intensive phase, a high proportion of PWTB in the intervention arm had gained 5% of their body weight (53.9% vs. 39.3%, p= 0.015). Similarly, at the end of treatment, a higher proportion of intervention arm participants gained 10% of their baseline body weight (55.8% vs. 41.0%, p=0.023). (Supplementary table 2).

**Figure 1:**
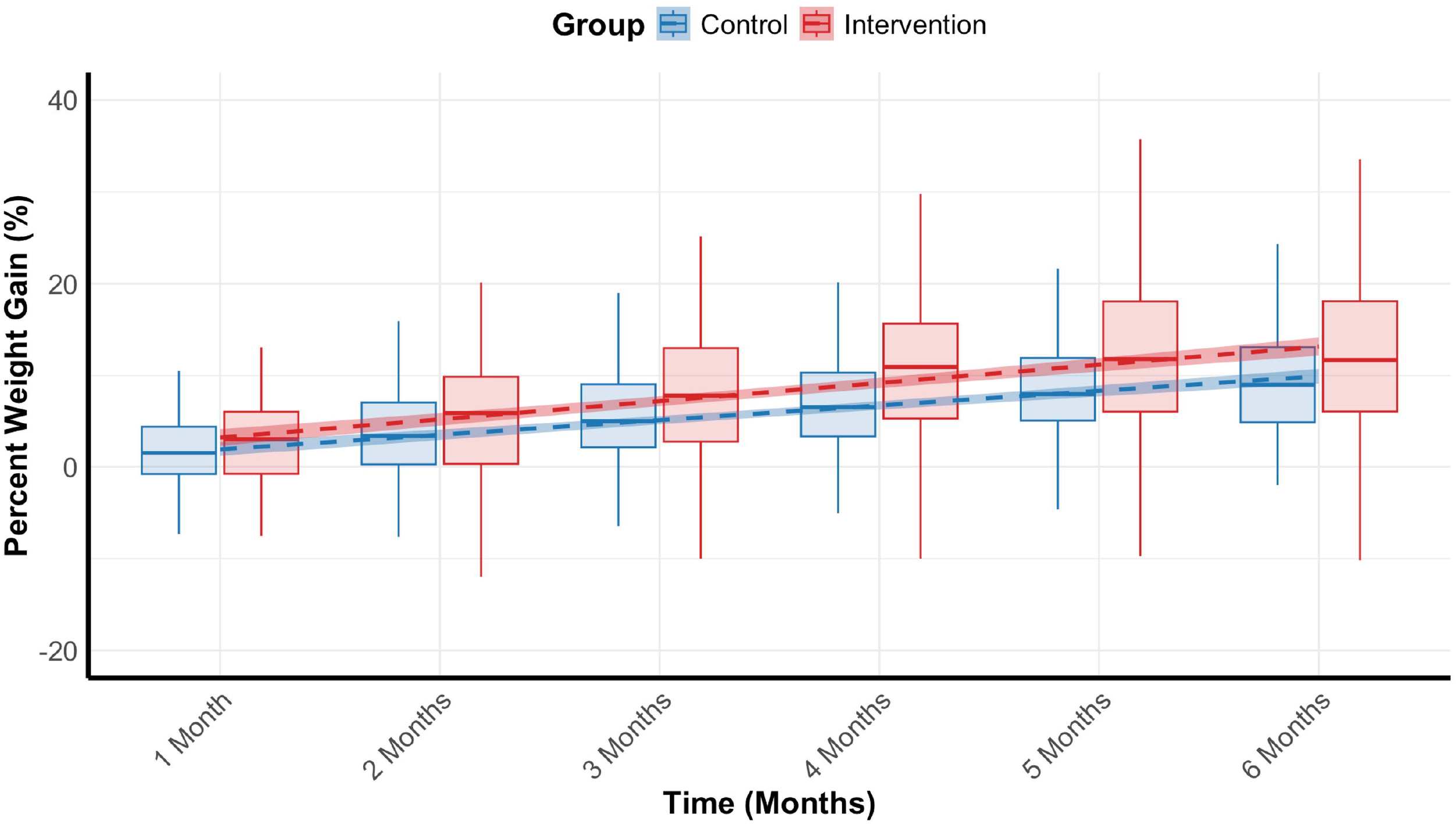
Boxplot comparing percentage of baseline body weight gained at different time points during TB treatment for intervention group (red) and control group (blue). Trend lines generated through linear regression.

### Primary outcomes

After adjusting for age, sex, baseline BMI, asset quintile, and alcohol use, intervention group participants had higher odds of 5% body weight gain at month 2 (adjust odds ratio [aOR]: 2.02 (95% confidence interval [CI]: 1.25-3.30). Intervention group participants also had higher odds of gaining 10% body weight by month 6 (aOR: 1.89 (95% CI: 1.13-3.18). (Supplementary table 3)

### Secondary outcomes

#### Acceptability and adherence

All participants (100%) found the sachet easy to open, though only 76.2% fully understood the instructions. Taste was acceptable to 96.7%, smell to 99.1%, and consistency to 99.9% of participants. Study personnel observed positive facial expressions in 76.2% of instances. Additionally, 89.9% of participants reported subjective health improvements after consuming EDNS. Adherence was high, with an overall rate of 88.7%, with 71% of participants never missing a sachet.

#### Adverse Events

Nausea and vomiting were more frequent in the intervention group, occurring in 10.2% vs. 4.3% (p = 0.055) and 7.5% vs. 3.6% (p = 0.149) of observed weeks, respectively. Abdominal fullness was also higher (12.9% vs. 5.4%, p = 0.032). Other adverse effects did not significantly differ between groups. (Supplementary table 4)

#### Hemoglobin recovery

Mean hemoglobin at baseline was 10.8 g/dL (SD 2.0). The hemoglobin gain at 2 months was 0.7 g/dL (SD 1.8) in the EDNS arm and 0.5 g/dL (SD 1.6) in the control arm (p=0.315), while at 6 months, it was 1.0 g/dL (SD 2.0) in the EDNS arm and 1.1 g/dL (SD 1.7) in the control arm (p=0.830).

## Discussion

To our knowledge, this is the largest randomized controlled trial to assess the impact of macronutrient support using a ready-to-use therapeutic food (RUTF) on weight gain among PWTB. We found that undernourished PWTB receiving EDNS gained more weight. Importantly, a higher proportion of EDNS recipients gained 5% body weight by the end of the intensive phase. This study provides a strong rationale for the use of EDNS to enhance the nutritional recovery of PWTB.

Notably, the weight gain among EDNS recipients was comparable to that seen among PWTB in the RATIONS study, which provided food baskets with 1200 Kcal per day. The RATIONS study PWTB had a mean baseline BMI of 16.4 (SD: 2.5) which was comparable to that of our participants. At month 2, 53.9% of EDNS recipients gained 5% of their baseline body weight, which was similar to the 54% of PWTB in RATIONS who attained the same metric. At the end of month 6, EDNS recipients had a median 11.7% weight gain (IQR: 6.1-18.1) which is almost identical to the RATIONS study (median 11.6%, IQR: 6.4-17.4). This comparison provides confidence that RUTFs such as EDNS are comparable to food baskets in improving weight gain.

One significant drawback of food baskets is the uncertainty about PWTB getting an adequate caloric load. Parents may instead prioritize their children’s nutrition, for instance. Gender dynamics may also come into play. Further, time-consuming preparation and fuel poverty may also inhibit the utilization of food baskets. And while Indian PWTB receive cash transfers through the Nikshay Poshan Yojana, the impact of cash transfers on tangible nutritional metrics among PWTB remains uncertain. RUTFs such as EDNS have been vetted in numerous clinical studies for use in contexts such as severe-acute malnutrition.^7^ Providing a dedicated nutritional supplement to persons with TB may help them attain a suitable caloric load, improving drug absorption, and potentially reducing drug toxicity, relapse, and mortality, particularly if provided promptly at treatment initiation.^8^ Moreover, the EDNS can be modified to meet the needs of special populations such as those with diabetes.

This study has several limitations. First, as an open-label trial, the lack of participant and investigator blinding may have introduced biases. Second, the study was conducted in two TB units in Faridabad which may limit generalizability to regions with different socio-economic or cultural contexts. Third, unmeasured factors such as dietary variations outside the intervention or social determinants of health could have influenced the outcomes. Finally, the exclusion of certain high-risk populations, such as those with diabetes or HIV, limits the applicability of our findings to these key and vulnerable subgroups. More research is needed to optimize the composition of EDNS for these populations as well as for children and pregnant women.

Our study demonstrates that EDNS effectively enhances weight gain among undernourished PWTB while being acceptable and with minimal adverse events, offering a practical and scalable solution for targeted caloric and micronutrient delivery. However, EDNS alone cannot resolve the widespread food insecurity faced by many PWTB. An integrated approach, as proposed by India’s National TB Elimination Program— combining cash transfers, food baskets, and EDNS--may provide a more comprehensive and sustainable strategy to improve nutritional outcomes and treatment success than any of these strategies.

## Data Availability

All data produced in the present study are available upon reasonable request to the authors

## Acknowledgement

We acknowledge the support provided by the study staff, investigators and participants in conduct of the study.

## References

1. Organization WH. Global tuberculosis report 2024. World Health Organization; 2024.

2. Sinha P, Davis J, Saag L, et al. Undernutrition and tuberculosis: public health implications. The Journal of infectious diseases. 2019;219(9):1356–1363.

3. Sinha P, Ponnuraja C, Gupte N, et al. Impact of Undernutrition on Tuberculosis Treatment Outcomes in India: A Multicenter Prospective Cohort Analysis. Clin Infect Dis. Nov 25 2022; doi:10.1093/cid/ciac915

4. Bhargava A, Bhargava M, Meher A, et al. Nutritional support for adult patients with microbiologically confirmed pulmonary tuberculosis: outcomes in a programmatic cohort nested within the RATIONS trial in Jharkhand, India. The Lancet Global Health. 2023;11(9):e1402–e1411.

5. Grobler L, Nagpal S, Sudarsanam TD, Sinclair D. Nutritional supplements for people being treated for active tuberculosis. Cochrane Database of Systematic Reviews. 2016/6// 2016;2016(6) doi:10.1002/14651858.CD006086.pub4

6. Kumar R, Krishnan A, Singh M, Singh UB, Singh A, Guleria R. Acceptability and Adherence to Peanut-Based Energy-Dense Nutritional Supplement Among Adult Malnourished Pulmonary Tuberculosis Patients in Ballabgarh Block of Haryana, India. Food and Nutrition Bulletin. 2020;41(4):438–445. doi:10.1177/0379572120952306

7. Schoonees A, Lombard MJ, Musekiwa A, Nel E, Volmink J. Ready-to-use therapeutic food (RUTF) for home-based nutritional rehabilitation of severe acute malnutrition in children from six months to five years of age. Cochrane Database of Systematic Reviews. 2019;(5)

8. Dauphinais MR, Koura KG, Narasimhan PB, et al. Nutritionally acquired immunodeficiency must be addressed with the same urgency as HIV to end tuberculosis. BMC Global and Public Health. 2024;2(1):1–9.

