## Supplementary materials for "Energy dense nutritional supplements improve weight gain among malnourished adults with drug-sensitive pulmonary tuberculosis: an open-label randomized controlled trial in Faridabad, India"

**Supplementary Figure 1:** Consort diagram

Assessed for eligibility (n= 729)

Excluded (n= 394)

- Not meeting inclusion criteria (n=269)
- Declined to participate (n=6)
- Excluded (n= 119)
  - Diabetes mellitus- 72, MDR-TB- 20, Pregnant or lactating women- 15, HIV sero-positive-6, too sick to participate=6

Randomized (n=335)

Intervention (n=171)

Control (n=164)

Lost to follow-up (n=8)

Death (n=8)

MDR detected at later stage(n=4)

(n=4)

Lost to follow-up (n=9)

Death (n=3)

MDR detected at later stage(n=2)

 Follow up at 2 months (n=150)

 Follow up at 2 months (n=151)

Lost to follow-up (n=9)

Return for 6-monthfollow-up (n=6)

Lost to follow-up (n=5)

Death (n=3)

Return for 6-month follow-up (n=1)

 Follow up at 6 months (n=143)

 Follow up at 6 months (n=148)

**Supplementary table 1:** Baseline characteristics of the study population

| **Variable** | **Control (n=164)** | **Intervention (n=171)** | **p-value** |
| --- | --- | --- | --- |
| **Age in years (mean (SD))** | 33.4 (14.7) | 33.1 (13.4) | 0.802 |
| **Male sex (%)** | 108 (65.9) | 91 (53.2) | 0.025 |
| **Asset Score quintile (%)**  1  2  3  4  5 | 33 (20.1)  33 (20.1)  33 (20.1)  37 (22.6)  28 (17.1) | 34 (19.9)  35 (20.5)  34 (19.9)  30 (17.5)  38 (22.2) | 0.701 |
| **Baseline BMI (kg/m2) (mean (SD))** | 16.4 (1.2) | 16.6 (1.5) | 0.250 |
| **Alcohol use (%)** | 12 (7.3) | 5 (2.9) | 0.114 |
| **Smoking (%)** | 71 (43.3) | 85 (49.7) | 0.286 |

BMI denotes body mass index; SD denotes standard deviation

**Supplementary table 2:** Nutritional outcomes

| **Variable** | **Control (n=164)** | **Intervention (n=171)** | **p-value** |
| --- | --- | --- | --- |
| Weight increase (kg) at 2 months, median (IQR) | 1.5 (2.8) | 2.4 (4.0) | 0.078 |
| Weight gain % at 2months, median (IQR) | 3.4 (6.8) | 5.9 (9.7) | 0.049 |
| 5% weight gain at 2 months, n(%) | 59 (39.3) | 82 (53.9) | 0.015 |
| Weight increase at 6 months, median (IQR) | 4 (3.7) | 4.9 (5.0) | 0.106 |
| Weight gain % at 6 months, median (IQR) | 9.0 (8.2) | 11.7 (12.0) | 0.047 |
| 10% weight gain at 6 months, n(%) | 55 (41.0) | 72 (55.8) | 0.023 |
| Attains BMI>18.5kg/m^2^, n (%) | 50 (37.3%) | 60 (46.5%) | 0.166 |

BMI denotes body mass index; IQR denotes interquartile ratio

**Supplementary table 3:** Regression analyses

| **Covariate** | **OR for 5% weight gain at 2 months (95% CI)** | **p-value** | **aOR for 5% weight gain at 2 months (95% CI)** | **p-value** |
| --- | --- | --- | --- | --- |
| EDNS | 1.81 (1.15-2.86) | 0.011* | 2.02 (1.25-3.30) | 0.005* |
| Age (years) | 1.00 (0.98-1.01) | 0.886 | 1.01 (0.99-1.02) | 0.504 |
| Male Sex | 0.70 (0.44-1.11) | 0.126 | 0.66 (0.38-1.12) | 0.125 |
| Asset Quintile  1  2  3  4  5 | Ref  0.69 (0.34-1.40)  0.71 (0.34-1.46)  0.62 (0.30-1.28)  0.35 (0.16-0.73) | 0.308  0.354  0.198  0.006* | Ref  0.64 (0.30-1.32)  0.72 (0.34-1.52)  0.62 (0.29-1.30)  0.30 (0.14-0.65) | 0.225  0.398  0.207  0.002* |
| Alcohol Use | 0.51 (0.17-1.40) | 0.200 | 0.36 (0.11-1.05) | 0.066 |
| Smoking | 1.17 (0.74-1.84) | 0.503 | - | - |
| Baseline BMI (kg/m2) | 0.95 (0.80-1.12) | 0.544 | 0.92 (0.77-1.10) | 0.362 |
| **Covariate** | **OR for 10% weight gain at 6 months (95% CI)** | **p-value** | **aOR for 10% weight gain at 6 months (95% CI)** | **p-value** |
| EDNS | 1.81 (1.11-2.97) | 0.017* | 1.89 (1.13-3.18) | 0.016* |
| Age (years) | 0.99 (0.98-1.01) | 0.538 | 1.00 (0.98-1.02) | 0.621 |
| Male Sex | 0.57 (0.34-0.94) | 0.027* | 0.57 (0.32-1.01) | 0.057 |
| Asset Quintile  1  2  3  4  5 | Ref  0.91 (0.43-1.94)  1.60 (0.76-3.45)  1.17 (0.54-2.55)  0.86 (0.39-1.91) | 0.810  0.221  0.698  0.719 | Ref  0.80 (0.36-1.76)  1.80 (0.83-3.99)  1.22 (0.54-2.76)  0.72 (0.31-1.65) | 0.580  0.141  0.639  0.438 |
| Alcohol Use | 2.24 (0.61-10.58) | 0.249 | 1.82 (0.46-9.07) | 0.418 |
| Smoking | 0.78 (0.48-1.27) | 0.319 | - | - |
| Baseline BMI (kg/m2) | 0.86 (0.71-1.03) | 0.104 | 0.86 (0.70-1.04) | 0.118 |

**Supplementary table 4:** Percentage of weeks with adverse effects

|  | **Percentage of weeks with adverse effects** | |  |
| --- | --- | --- | --- |
| **Symptom** | **Control group** | **Intervention group** | **p-value** |
| Nausea | 4.3 | 10.2 | <0.001 |
| Vomiting | 3.6 | 7.5 | 0.003 |
| Diarrhea | 0.1 | 0.4 | 0.081 |
| Abdominal pain | 3.9 | 5.1 | 0.282 |
| Chills | 1.2 | 1.6 | 0.392 |
| Headache | 6.0 | 5.1 | 0.4 |
| Bodyache | 19.5 | 18.3 | 0.627 |
| Wheeze | 2.9 | 2.7 | 0.827 |
| Irritability | 1.8 | 1.6 | 0.832 |
| Allergy | 0.7 | 1.1 | 0.397 |
| Bloating | 5.4 | 12.9 | <0.001 |

p-value calculated using student’s t-test
